# Factors associated with rabies vaccination uptake among dog owners in Butaleja Town Council, Eastern Uganda: A cross-sectional study

**DOI:** 10.64898/2026.08.14.26360440

**Authors:** Salim Hasahya, Saul Kamukama, Stella Maris Lunkuse, David Lubogo

## Abstract

Rabies remains a major zoonotic public health threat, associated with approximately 59,000 annual deaths globally, with Africa bearing 36% of this burden. In Uganda, an estimated 36 human deaths and 1.8 million dog bite exposures occur annually, yet national dog vaccination coverage remains critically low at approximately 10%. This study investigated factors associated with rabies vaccination uptake among dog owners in Butaleja Town Council, Eastern Uganda, a high-risk setting. A cross-sectional survey was conducted among 173 dog owners between May 18 and June 28, 2024, using semi-structured questionnaires. The primary outcome was vaccination uptake (≥1 dose in the past year). Modified Poisson regression with robust variance estimated adjusted prevalence ratios for all socio-demographic, veterinary system, health system, dog-related, and knowledge factors. Among 173 dog owners, 57.2% vaccinated at least one dog, corresponding to 54.6% (166/304) of all dogs vaccinated. Factors associated with vaccination uptake included older age (21–40 years: aPR 1.43, 95% CI: 1.15–1.71; 41–60 years: aPR 1.53, 95% CI: 1.22–1.82; ≥61 years: aPR 1.56, 95% CI: 1.33–2.23), higher education (primary: aPR 1.20, 95% CI: 1.00–1.45; secondary: aPR 1.55, 95% CI: 1.20–2.00), higher income, access to veterinary clinics (aPR 2.50, 95% CI: 1.52–3.75), and participation in community education sessions (aPR 1.71, 95% CI: 1.34–2.15). While household-level uptake showed moderate engagement, the resulting dog population vaccination coverage of 48.4% remained substantially below the 70% World Health Organization threshold required for herd immunity. Achieving the “Zero by 30” elimination dog-bite mediated Rabies target in this setting requires targeted interventions addressing the identified associated factors including; enhanced community education, improved veterinary service accessibility, reminder systems, and strategies to convert partial household vaccination into complete coverage particularly among younger, less educated, and lower-income dog owners.

**Author Summary:** Rabies remains a neglected tropical disease that claims approximately 59,000 lives annually, predominantly in Africa and Asia. While mass dog vaccination is the most effective control strategy, many communities struggle to reach the 70% coverage needed to mitigate rabies transmission. This study investigated why dog owners in a high-risk Ugandan community do or do not vaccinate their animals.

We surveyed 173 dog owners and found an important gap; while 57% of owners had vaccinated at least one dog, only 48% of all dogs were actually vaccinated far below the global target. This distinction matters because it shows that even owners who participate in vaccination programs often leave some dogs unprotected, undermining efforts to achieve herd immunity.

Several factors were associated with vaccination. Owners with better access to veterinary clinics, knowledge of where to get vaccines, and participation in community education were more likely to vaccinate. Receiving vaccination reminders also made a significant difference. Conversely, limited awareness, poor service accessibility, and socioeconomic challenges prevented many from vaccinating their dogs.

These findings point to practical solutions: improving communication about vaccination locations, sending reminders, and strengthening community education could help more families fully vaccinate all their dogs. For Uganda and similar settings, addressing these barriers is essential to reach the global goal of eliminating rabies deaths by 2030.

## 1.0 Introduction

Rabies is a fatal zoonotic viral disease that affects all warm-blooded mammals, including humans. It is primarily transmitted through the saliva of infected animals, especially dogs, and causes acute encephalitis that is nearly always fatal once symptoms appear (1). Globally, rabies causes an estimated 59,000 human deaths annually, with Africa accounting for over 60% of this burden due to limited access to preventive measures and weak surveillance systems (2, 3).

In Africa, domestic dogs are responsible for approximately 98% of human rabies cases, making dog-mediated transmission the primary concern for public health interventions (2, 4). Uganda is among the countries with a high rabies burden, recording an estimated 133 human rabies deaths per year and over 12,234 post-exposure prophylaxis (PEP) treatments annually (3). Between 2015 and 2020, Ministry of Health reported 36 annual fatalities and 1.8 million animal bite exposures, underscoring the persistent threat of rabies across the country (5). The economic impact is substantial, with annual PEP costs estimated at US$ 7.8 million, equivalent to 0.03% of Uganda’s GDP (6).

Despite the availability of effective rabies vaccines through district veterinary services under the guidance of Uganda’s Ministry of Agriculture, Animal Industry and Fisheries (MAAIF), rabies vaccination uptake by dog owners remains critically low in Uganda. While national coverage is estimated at 10%, uptake is hindered by factors such as limited awareness, affordability, accessibility, and misconceptions about rabies and its prevention (7). These behavioral and systemic barriers pose a significant challenge to achieving World Health Organization’s recommended 70% coverage for herd immunity (8).

Uganda has adopted the Stepwise Approach towards Rabies Elimination (SARE), scoring 0.5 out of 5, indicating early-stage implementation with situational data collection but limited intervention rollout (3). Butaleja Town Council (TC), located in Butaleja District, Eastern Uganda, is a high-risk area in a district reporting the second-highest incidence of suspected rabies cases in the Bukedi sub-region (9–11). With an estimated dog population of 793, vaccination coverage remains undocumented and presumed low, perpetuating the risk of human exposure. This study examined the factors associated with rabies vaccination uptake among dog owners in Butaleja TC using a cross-sectional design. Findings aim to inform targeted interventions and support Uganda’s goal of eliminating human rabies deaths by 2030 (2).

## 2.0 Methods

### 2.1 Study area and population

The study was conducted in Butaleja TC, part of Butaleja District in Eastern Uganda. Butaleja TC has an estimated population of 27,500 and comprises six parishes (Nanyulu, Butaleja, Lujjehe, Sagenda, Bunghaji, and Hisega) and 29 villages (12). The district was established on July 1, 2005, and carved out of Tororo District (10). The study population consisted of dog owners residing in Butaleja TC, who were recruited and interviewed between May 18th and June 28th, 2024. Dog owners were selected as the primary decision-makers regarding rabies vaccination for their dogs, providing insights into factors associated with rabies vaccination uptake.

### 2.2 Study design and sample size

A cross-sectional study design was employed to assess factors associated with rabies vaccination uptake among dog owners in Butaleja TC. The target population consisted of 312 dog owners, as determined from records maintained by the Butaleja district production department under Veterinary section. The sample size was calculated using the formula for finite populations (13).

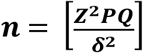

Where: n = sample size

Z = z-score for a 95% confidence level (1.96)

P = since there’s no studies for P, we used P = 0.5 as a conservative estimate

δ = is the desired margin of error (0.05 for a 5% margin of error)

We substituted the values into the formula.

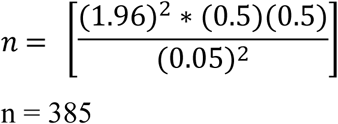

n = 385

Since the population size is finite (312), the adjusted sample size formula to be used was:

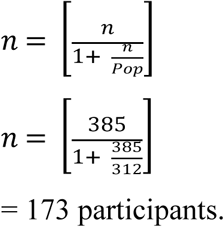

= 173 participants.

### 2.3 Sampling procedure

The participants were selected through simple random sampling from a list of dog owners obtained from LC1 records, with each village’s dog owners assigned unique identifiers to ensure proportional representation across all villages using a random number generator, thereby giving every dog owner an equal chance of being included in the study (14). To get to the households with dog owners to interview, the sampling procedure involved obtaining village lists, along with the number of dogs and dog owners per village, from Butaleja District production office records, followed by generating a verified list of dog owners by contacting LC1 chairpersons to confirm ownership. The total number of dog owners in Butaleja TC was then calculated by aggregating the figures from all the six parishes of Butaleja TC, after which a comprehensive household list was compiled, with each dog owning household assigned a unique identifier for random sampling. Selected participants were then reached out to through direct visits by the research team, coordinated in advance with the respective LC1 chairpersons to ensure accessibility and consent. This systematic approach ensured an unbiased and representative selection of participants.

#### 2.3.1 Inclusion and exclusion criteria

Inclusion criteria required participants to be residents of Butaleja TC for at least one year, own at least one dog for a minimum of one year, acting as the primary decision-maker for rabies vaccination, and be aged 18 years or older. Exclusion criteria applied to individuals exhibiting significant emotional distress or cognitive impairment, assessed using the Kessler Psychological Distress Scale (K10), with a score ≥30 indicating exclusion (15, 16).

### 2.4 Data collection and variables

Data were collected using semi-structured questionnaires administered by trained research assistants. The questionnaire was adapted from validated tools (17, 18) and was pretested from Nabiganda TC also in Butaleja district for clarity and relevance (19, 20).

The dependent variable was dog owners’ rabies vaccination uptake, measured as a binary outcome indicating whether a dog owner had a dog vaccinated received at least one rabies vaccine dose within the past year (1, 21, 22). This measurement captured the proportion of dog owners in Butaleja TC whose dogs were vaccinated, offering insight into current rabies control status. Independent variables were measured across predisposing, enabling, and needs domains to comprehensively assess factors associated with dog owners’ vaccination uptake (23). Predisposing factors included socio-demographic characteristics; age, sex, education, income, occupation, religion, marital status, and family size, collected through administering semi-structured questionnaires. Enabling factors assessed access to PEP, awareness, disease recognition, community engagement, health promotion, public awareness, service accessibility, vaccination availability, and veterinary service use. Needs factors measured knowledge and practices related to rabies transmission, prevention, social norms, and cultural influences, alongside dog-related characteristics such as age, sex, confinement status, vaccination history, breed, and ownership purpose. All variables were assessed using rigorously pretested semi-structured questionnaires adapted from validated tools (17, 18).

### 2.5 Data preparation and descriptive analysis of dog owners’ and their dogs

#### 2.5.1 Data management

The data management process prioritized the security, integrity, and confidentiality of all participant information. Survey data were backed up on Google Drive, and a copy was used for subsequent analyses. Data cleaning was conducted in MS Excel before being exported to Stata V14.0. The outcome variable, dog owners’ rabies vaccination uptake, was assessed as a binary measure and summarized at two levels: Yes, for vaccinated and No for none vaccinated, expressed as a proportion (24, 25).

#### 2.5.2 Data analysis

The data analysis process aimed to extract meaningful insights from the collected data to understand factors associated with rabies vaccination uptake among dog owners in Butaleja TC. Survey-data-restricted Modified Poisson regression was applied to assess factors associated with uptake of dog rabies vaccination, with associations measured as prevalence ratios (PRs) and their 95% confidence intervals (26, 27). This approach is appropriate for binary outcomes in cross-sectional survey data because it incorporates survey design restrictions, uses robust variance estimation, and yields directly interpretable PRs without overstating associations.

At bivariate analysis, variables with P<0.25 were retained for multivariate analysis. The multivariate model assessed the joint association between all selected independent variables and the outcome, with significance considered at P<0.05 (28). A chunk test was used to identify interaction terms, and confounding was evaluated only in the absence of interaction by assessing a ≥10% change in the effect measure in the presence of a third variable (29). These procedures allowed explicit identification of interacting variables and confounders within the final model.

A correlation matrix was further generated to determine whether independent variables were correlated. Where pairs of variables showed correlation coefficients ≥0.4, one variable was removed from subsequent analyses to prevent multicollinearity (30). This ensured model stability and avoided biased estimates. The correlation analysis revealed several variables with moderate to strong correlations. Specifically, education level and income showed a correlation coefficient of 0.52, indicating that higher educational attainment was associated with higher income levels. Additionally, access to veterinary clinics and awareness of vaccination locations were correlated at 0.45, suggesting that owners with better access to veterinary services were also more likely to know where vaccination services were offered. Knowledge of rabies transmission and knowledge of rabies signs demonstrated a correlation of 0.48, reflecting the interrelated nature of rabies-related knowledge domains. To address these correlations and prevent multicollinearity, the following variables were removed from the multivariate model: income (due to correlation with education), awareness of vaccination locations (due to correlation with access to veterinary clinics), and knowledge of rabies signs (due to correlation with knowledge of rabies transmission). Education, access to veterinary clinics, and knowledge of rabies transmission were retained in the final model based on their stronger theoretical relevance to vaccination uptake and their lower correlation with other retained variables.

Model adequacy was assessed using Akaike’s Information Criterion (AIC) and Bayesian Information Criterion (BIC) (31). The model with the lowest AIC and BIC values was considered the best fitting (32, 33). The final multivariate model demonstrated adequate fit with AIC = 342.1 and BIC = 398.7, indicating good parsimony and explanatory power.

### 2.6 Quality assurance

Quality assurance procedures were implemented throughout the study to ensure the accuracy, completeness, consistency, and reliability of the data. Research assistants were trained on the study objectives, questionnaire administration, ethical requirements, confidentiality, and culturally appropriate interviewing techniques. Role-play and practical exercises were used to standardize data-collection procedures and improve interviewer consistency (34).

Before the main data collection, the questionnaire was pretested among eligible dog owners in Nabiganda Town Council, a setting outside the study area, to assess its clarity, relevance, comprehensibility, and suitability for the study population. Identified ambiguities, inconsistencies, and gaps were revised before the final questionnaire was administered.

During data collection, supervisors conducted daily quality checks of completed questionnaires to identify missing responses, inconsistencies, and recording errors. Identified discrepancies were resolved promptly through discussion with the data collectors and, where necessary, follow-up with participants. Reported dog vaccination status was cross-checked against available vaccination certificates and veterinary records, where applicable, to improve the accuracy of vaccination-status classification.

Following data collection, questionnaires were reviewed for completeness, coded, cleaned, and checked for consistency before analysis. Data were securely stored with access restricted to authorized members of the research team. Missing data were assessed before analysis, and where imputation was necessary, the prespecified approach was applied consistently. These procedures enhanced the validity, completeness, accuracy, and reliability of the study dataset.

### 2.7 Ethical considerations

Prior to data collection, ethical approval was obtained from the Makerere University Institutional Review Board, with the Makerere School of Public Health Research and Ethics Committee (MakSPH-REC) assigning study protocol number 397, ensuring full compliance with established guidelines. Research assistants were trained to uphold core ethical principles throughout the study, including informed consent, voluntary participation (both verbal and written consent was obtained from the participants before data collection), respect for participants, and strict confidentiality. To protect privacy, participant data were anonymized and stored securely on password-protected systems accessible only to the research team, in line with recommended practices for safeguarding sensitive information (35, 36). The study posed minimal risk and was designed to maximize participant benefit while maintaining the highest ethical standards throughout.

## 3.0 Results

### 3.1 Socio-demographic characteristics of respondents

The study surveyed 173 dog owners, achieving 100% response rate. The age distribution of the respondents was varied, with the majority falling between 21 and 60 years.

The majority of dog owners were male, comprising 67.1% of the sample, while females accounted for 32.9%. Educational attainment among dog owners showed that a significant proportion had only primary education (68.2%). Those with secondary education made up 19.7%, and a smaller percentage had attained post-secondary education (6.9%).

The vast majority of respondents were farmers, making up 80.3% of the participants. This was followed by those involved in business or trade (6.4%), civil servants (2.9%), and the unemployed (3.5%). A small segment of the respondents fell into other occupations (6.9%), refer to **Table 1**.

**Table 1:** Socio-demographic characteristics of interviewed dog owners’(N=173)

| Factor | Attribute | Frequency | Percent (%) |
| --- | --- | --- | --- |
| Age | ≤20 | 15 | 8.7 |
|  | 21-40 | 66 | 38.2 |
|  | 41-60 | 61 | 35.3 |
|  | ≥61 | 31 | 17.9 |
| Sex | Male | 116 | 67.1 |
|  | Female | 57 | 32.9 |
| Education | None | 9 | 5.2 |
|  | Primary | 118 | 68.2 |
|  | Secondary | 34 | 19.7 |
|  | Post-secondary | 12 | 6.9 |
| Occupation | Farmer | 139 | 80.3 |
|  | Business/trade | 11 | 6.4 |
|  | Civil servant | 5 | 2.9 |
|  | Unemployed | 6 | 3.5 |
|  | Other | 12 | 6.9 |
| Religion | Catholic | 18 | 10.4 |
|  | Islam | 23 | 13.3 |
|  | Pentecostal | 11 | 6.4 |
|  | Protestant | 121 | 69.9 |
| Marital status | Single | 20 | 11.6 |
|  | Married | 142 | 82.1 |
|  | Widowed | 11 | 6.4 |
| Type of family | Nuclear | 77 | 44.5 |
|  | Extended | 96 | 55.5 |
| Monthly income | ≤ 200,000 | 124 | 71.7 |
|  | 210,000 - 400,000 | 28 | 16.2 |
|  | ≥ 400,001 | 21 | 12.1 |

#### 3.1.1 Characteristics of dogs owned by interviewed dog owners

As shown in **Table 2**, the majority of the dogs were male, accounting for 73.4% of the sample, while females made up 26.6%. Most dogs roamed freely, representing 85.8% of the population. A smaller proportion was confined mostly indoors (8.6%) or outdoors (5.6%).

**Table 2:** Characteristics of dogs in surveyed households.

| Variable | Attribute | Frequency | Percent (%) |
| --- | --- | --- | --- |
| Sex | Male | 223 | 73.4 |
|  | Female | 81 | 26.6 |
| Confinement status | Confined mostly indoors | 26 | 8.6 |
|  | Confined mostly outdoors | 17 | 5.6 |
|  | Roams freely | 261 | 85.8 |
| Breed | Local | 265 | 87.1 |
|  | Mixed | 39 | 12.9 |
| Dog origin | Born in this town | 267 | 87.8 |
|  | Brought from another location | 37 | 12.2 |
| Purpose of being kept | Security | 281 | 92.4 |
|  | Hunting | 23 | 7.6 |
| Household Dog ownership | One | 161 | 53 |
|  | Two | 56 | 18.4 |
|  | More than three | 87 | 28.6 |

The breed composition indicated a strong preference for local breeds, with 87.1% of the dogs being local and 12.9% being mixed breeds. In terms of origin, 87.8% of the dogs were born in the same town, while 12.2% were brought from other locations. The primary purpose for keeping these dogs was predominantly for security, with 92.4% of the dogs kept for this reason, while 7.6% were kept for hunting.

Household dog ownership varied, with 53.0% of households owning one dog, 18.4% owning two dogs, and 28.6% having more than three dogs.

***Table 3*** shows that 57.2% of dog owners vaccinated at least one dog, with 81.8% of these owners vaccinating all their dogs. Overall, 48.4% of the total dog population was vaccinated, indicating moderate uptake at the household level.

**Table 3:** Rabies vaccination coverage from owner-level effort to full household compliance and overall population protection among dog-owning households in Butaleja TC.

| Tier | Level of analysis | Category | Numerator | Denominator | % |
| --- | --- | --- | --- | --- | --- |
| 1 | Dog ownership level | Dog owners that vaccinated at least one of their dogs | 99 | 173 | 57.2 |
| 2 | Dog owners' compliance level | Out of Dog owners' that vaccinated, those that vaccinated ALL of their dogs | 81 | 99 | 81.8 |
|  |  | Dog owners' that vaccinated ONLY SOME of their dogs. | 18 | 99 | 18.2 |
| 3 | Dog vaccination prevalence | Total dogs that were actually vaccinated | 147 | 304 | 48.4 |

### 3.2 Rabies vaccination uptake among dog owners in Butaleja TC

The study revealed that out of 173 respondents, 99 (57.2%) had at least vaccinated one of their dogs against rabies, while 74 (42.77%) had not. Slightly more than half of the dog owners ensured their dogs received the vaccination, whereas nearly 42% did not (refer **Table 4**)

**Table 4:** Rabies vaccination uptake among surveyed dog owners.

| Vaccination Status of their dogs | Frequency | Percent (%) |
| --- | --- | --- |
| Vaccinated | 99 | 57.2 |
| Not vaccinated | 74 | 42.8 |
| Total | 173 | 100.0 |

Among the 99 dog owners’ who did vaccinate their dogs, a significant majority (81.8%) achieved complete vaccination coverage for all their dogs, while a smaller proportion (18.2%) did not achieve full vaccination coverage (as indicated in **Table 4 Table 5**)

**Table 5:** Proportion of dog owners’ who achieved full vaccination uptake for Rabies vaccine.

| Dogs owned | Dog owners' who vaccinated all | Dog owners who didn't vaccinate all | Total |
| --- | --- | --- | --- |
| 1 | 49 | 0 | 49 |
| 2 | 18 | 2 | 20 |
| 3 | 9 | 6 | 15 |
| 4 | 4 | 4 | 8 |
| 5 | 0 | 6 | 6 |
| 6 | 1 | 0 | 1 |
|  | 81 (81.8%) | 18 (18.2%) | 99 (100%) |

### 3.3 Factors associated with Rabies vaccination uptake among dog owners’ in Butaleja TC. Socio-demographic factors

After adjustment for potential confounders, age was significantly associated with rabies vaccination uptake among dog owners. Compared with owners aged ≤20 years, those aged 21–40 years (aPR = 1.43; 95% CI: 1.15–1.71), 41–60 years (aPR = 1.53; 95% CI: 1.22–1.82), and ≥61 years (aPR = 1.56; 95% CI: 1.33–2.23) were significantly more likely to have their dogs vaccinated.

Sex was also significantly associated with vaccination uptake. Female dog owners were less likely to vaccinate their dogs compared with male owners (aPR = 0.78; 95% CI: 0.65–0.95).

Educational attainment showed a positive association with vaccination uptake. Dog owners with primary education (aPR = 1.20; 95% CI: 1.00–1.45) and secondary education (aPR = 1.55; 95% CI: 1.20–2.00) were more likely to vaccinate their dogs compared with those without formal education, while post-secondary education was not significantly associated with vaccination uptake.

Religious affiliation was significantly associated with rabies vaccination uptake. Protestant (aPR = 1.26; 95% CI: 1.12–1.63) and Pentecostal (aPR = 1.25; 95% CI: 1.15–1.82) dog owners were more likely to vaccinate their dogs compared with Catholics, whereas no significant difference was observed among Muslim dog owners.

Marital status was independently associated with vaccination uptake. Married (aPR = 1.25; 95% CI: 1.14–1.33) and widowed dog owners (aPR = 1.61; 95% CI: 1.28–1.98) were more likely to vaccinate their dogs compared with single owners.

Household structure was also significant, with dog owners living in extended families being more likely to vaccinate their dogs than those in nuclear families (aPR = 1.45; 95% CI: 1.33–1.83). In addition, higher household income was associated with increased vaccination uptake, particularly among owners earning ≥400,001 UGX per month (aPR = 1.35; 95% CI: 1.05–1.75) compared with those earning ≤200,000 UGX (as indicated in **Table 6**)

**Table 6:** Socio-demographic, veterinary system, Health system and Dog-related factors associated with Rabies vaccination uptake among dog owners.

| Variable | Vaccinated<br>n(%) | None vaccinated<br>n(%) | cPR(95% CI) | aPR (95% CI) |
| --- | --- | --- | --- | --- |
| **Age** |  |  |  |  |
| $\leq 20$ | 6 (40.0) | 9(60.0) | 1 | 1 |
| 21-40 | 39 (59.1) | 27 (40.9) | 1.31 (1.20 - 1.88) | 1.43 (1.15 - 1.71) *** |
| 41-60 | 36 (59.0) | 25 (41.0) | 1.46 (1.34 - 2.04) | 1.53 (1.22 - 1.82) *** |
| $\geq 61$ | 19 (61.3) | 12 (38.7) | 1.51 (1.40 - 2.03) | 1.56 (1.33 - 2.23) *** |
| **Sex** |  |  |  |  |
| Male | 72 (62.1) | 44 (37.9) | 1 | 1 |
| Female | 28 (49.1) | 29 (50.9) | 0.75 (0.60 - 0.90) | 0.78 (0.65 - 0.95) ** |
| **Education** |  |  |  |  |
| None | 5 (55.6) | 4 (44.4) | 1 | 1 |
| Primary | 67 (56.8) | 51 (43.2) | 1.25 (1.05 - 1.48) | 1.20 (1.00 - 1.45) * |
| Secondary | 22 (64.7) | 12 (35.3) | 1.60 (1.25 - 2.05) | 1.55 (1.20 - 2.00) *** |
| Post-secondary | 6 (50.0) | 6 (50.0) | 1.30 (0.70 - 1.43) | 1.05 (0.72 - 1.50) * |
| **Religion** |  |  |  |  |
| Catholic | 9 (50.0) | 9 (50.0) | 1 | 1 |
| Islam | 11 (47.8) | 12 (52.2) | 1.06 (0.70 - 1.60) | 1.05 (0.68 - 1.55) |
| Pentecostal | 7 (63.6) | 4 (36.4) | 1.27 (1.07 - 1.85) | 1.25 (1.15 - 1.82) ** |
| Protestant | 73 (60.3) | 48 (39.7) | 1.26 (1.05 - 1.66) | 1.26 (1.12 - 1.63) *** |
| **Marital status** |  |  |  |  |
| Single | 10 (50.0) | 10 (50.0) | 1 | 1 |
| Married | 86 (60.6) | 56 (39.4) | 1.06 (0.85 - 1.33) | 1.25 (1.14 - 1.33) *** |
| Widowed | 4 (36.4) | 7 (63.6) | 0.54 (0.25 - 1.17) | 1.61 (1.28 - 1.98) *** |
| **Type of family** |  |  |  |  |
| Nuclear | 43 (55.8) | 34 (44.2) | 1 | 1 |
| Extended | 57 (59.4) | 39 (40.6) | 1.04 (1.02 - 1.32) | 1.45 (1.33 - 1.83) *** |
| **Monthly income** |  |  |  |  |
| ≤ 200,000 | 67 (54.0) | 57 (46.0) | 1 | 1 |
| 200,001 - 400,000 | 16 (57.1) | 12 (42.9) | 0.94 (0.66 - 1.34) | 1.05 (1.01 - 1.35) * |
| ≥ 400,001 | 17 (81.0) | 4 (19.0) | 1.39 (1.08 - 1.79) | 1.35 (1.05 - 1.75) ** |
| **Policy awareness** |  |  |  |  |
| Yes | 69 (57.9) | 50 (42.1) | 1 | 1 |
| No | 29 (53.7) | 25 (46.3) | 0.43 (0.38, 0.93) | 0.33 (0.13 - 0.98)*** |
| **Vaccination reminders** |  |  |  |  |
| Yes | 85 (64.3) | 47 (35.7) | 1 | 1 |
| No | 13 (31.7) | 28 (68.3) | 0.20 (0.12, 0.33) | 0.22 (0.13 - 0.35)** |
| **Vet clinic access** |  |  |  |  |
| No | 4 (21.1) | 15 (78.9) | 1 | 1 |
| Yes | 93 (60.4) | 61 (39.6) | 2.60 (1.01, 3.84) | 2.50 (1.52 - 3.75)** |
| **Aware of vaccination location** |  |  |  |  |
| No | 6 (15.8) | 32 (84.2) | 1 | 1 |
| Yes | 91 (67.4) | 44 (32.6) | 1.67 (1.31 - 2.02) | 1.41 (1.33 - 2.12)** |
| **Aware of PEP** |  |  |  |  |
| No | 15 (54.6) | 13 (46.4) | 1 | 1 |
| Yes | 83 (57.2) | 62 (42.8) | 1.05 (1.01 - 1.27) | 1.08 (1.06 - 1.72)** |
| **Participation in community education sessions** |  |  |  |  |
| No | 64 (48.9) | 67 (51.1) | 1 | 1 |
| Yes | 36 (85.7) | 6 (14.3) | 1.75 (1.38 - 2.22) | 1.71 (1.34 - 2.15)** |
| **Comfort discussing dog health** |  |  |  |  |
| No | 8 (38.1) | 13 (61.9) | 1 | 1 |
| Yes | 90 (59.2) | 62 (40.8) | 1.46 (1.22 - 1.87) | 1.53 (1.25 - 2.65)** |
| **Confinement status** |  |  |  |  |
| Confined mostly indoors | 2 (33.3) | 4 (66.7) | 1 | 1 |
| Confined mostly outdoors | 6 (100.0) | 0 (0.0) | 1.51 (1.42 - 1.83) | 2.31 (1.62 - 2.01)** |
| Roams freely | 92 (57.) | 69 (42.9) | 1.73 (1.50 - 2.20) | 1.54 (1.40 - 1.74)** |
| **Breed** |  |  |  |  |
| Mixed | 4 (80.0) | 1 (20.0) | 1 | 1 |
| Local | 96 (57.1) | 72 (42.9) | 1.33 (1.05 - 1.85) | 1.30 (1.13 - 1.80)** |
| **Dog origin** |  |  |  |  |
| Born in this town | 92 (60.1) | 61 (39.9) | 1 | 1 |
| Brought from another location | 8 (40.0) | 12 (60.0) | 0.66 (0.38 - 0.84) | 0.52 (0.40 - 0.76)*** |
| **Purpose of being kept** |  |  |  |  |
| Hunting | 2 (50.0) | 2 (50.0) | 1 | 1 |
| Security | 98 (58.0) | 71 (42.0) | 1.20 (1.02 - 1.50) | 1.25 (1.05 - 1.60)** |
| **Dog number** |  |  |  |  |
| One | 46 (50.5) | 45 (49.5) | 1 | 1 |
| Two | 23 (60.5) | 15 (39.5) | 1.10 (0.78 - 1.55) | 1.12 (1.05 - 1.60)** |
| More than three | 32 (72.7) | 12 (27.3) | 1.42 (1.10 - 1.82) | 1.45 (1.12 - 1.90)*** |

## Veterinary system factors

After adjustment for potential confounders, several veterinary systems–related factors were significantly associated with rabies vaccination uptake among dog owners in Butaleja TC( **Table 6**). Dog owners who were not aware of existing dog vaccination policies had a significantly lower prevalence of vaccinating their dogs compared with those who were aware (aPR = 0.33; 95% CI: 0.13–0.98). Similarly, owners who did not receive vaccination reminders were substantially less likely to vaccinate their dogs than those who received reminders (aPR = 0.22; 95% CI: 0.13–0.35).

Access to veterinary services showed a strong positive association with vaccination uptake. Dog owners with access to veterinary clinics were more than twice as likely to vaccinate their dogs compared to those without access (aPR = 2.50; 95% CI: 1.52–3.75). Awareness of vaccination locations was also independently associated with higher vaccination prevalence (aPR = 1.41; 95% CI: 1.33–2.12).

Furthermore, dog owners who were aware of post-exposure prophylaxis (PEP) (aPR = 1.08; 95% CI: 1.06–1.72), had participated in community education sessions (aPR = 1.71; 95% CI: 1.34– 2.15), and felt comfortable discussing dog health issues with veterinary personnel (aPR = 1.53; 95% CI: 1.25–2.65) demonstrated significantly higher uptake of rabies vaccination (Refer to **Table 6**).

## Health system factors

Dog owners who were aware of the vaccination location were more likely to vaccinate their dogs (aPR: 1.41, 95% CI: 1.33 - 2.12) compared to those who were not aware. Likewise, respondents who were aware of PEP were slightly more likely to vaccinate their dogs (aPR: 1.08, 95% CI: 1.06 - 1.72) compared to those who were not aware. Participation in community education sessions had a significant association with vaccination uptake. Owners who participated in these sessions were more likely to vaccinate their dogs (aPR: 1.71, 95% CI: 1.34 - 2.15) compared to those who did not participate. Owners who were comfortable discussing dog health were significantly more likely to vaccinate their dogs (aPR: 1.53, 95% CI: 1.25 - 2.65) compared to those who were not comfortable **Table 6**.

## Dog related factors

Confinement status was significantly associated with rabies vaccination rates. Dogs confined mostly outdoors had a higher likelihood of being vaccinated (aPR: 2.31, 95% CI: 1.62 - 2.01) compared to those confined mostly indoors. Similarly, dogs that roamed freely were more likely to be vaccinated (aPR: 1.54, 95% CI: 1.40 - 1.74) compared to those confined indoors. Dog origin also played a role—dogs born in the town were more likely to be vaccinated compared to those brought from another location, with a lower likelihood of vaccination for the latter group (aPR: 0.52, 95% CI: 0.40 - 0.76).

The purpose for which dogs were kept influenced their vaccination status. Dogs kept for security purposes had a higher likelihood of being vaccinated (aPR: 1.25, 95% CI: 1.05 - 1.60) compared to those kept for hunting.

The number of dogs owned by the household was another significant factor. Households with two dogs were more likely to vaccinate their dogs (aPR: 1.12, 95% CI: 1.05 - 1.60) compared to households with only one dog. Furthermore, households with more than three dogs showed an even higher likelihood of vaccinating their dogs (aPR: 1.45, 95% CI: 1.12 - 1.90) compared to those with a single dog (as indicated in **Table 6**).

## Influence of knowledge of dog owners on Rabies vaccination uptake

Regarding the mode of transmission, respondents who answered two questions correctly were more likely to vaccinate their dogs (aPR: 1.43, 95% CI: 1.32 - 2.44) compared to those who provided no correct answers. Those with four or more correct answers also showed higher vaccination rates (aPR: 1.05, 95% CI: 1.01 - 1.35) compared to those with zero correct answer.

Knowledge about the importance of vaccination had a significant positive impact. Respondents who answered two questions correctly were significantly more likely to vaccinate their dogs (aPR: 1.85, 95% CI: 1.23 - 2.41). Those who answered three or more questions correctly were even more likely to vaccinate their dogs (aPR: 2.05, 95% CI: 1.10 - 3.80) compared to those who didn’t know any correct answer. Awareness of rabies signs also played a crucial role. Respondents who answered two questions correctly about rabies signs had a higher likelihood of vaccinating their dogs (aPR: 1.42, 95% CI: 1.21 - 2.51). The likelihood increased further for those who answered three questions correctly (aPR: 1.73, 95% CI: 1.22 - 2.52) compared to those who didn’t know any correct answer (as indicated in **Table 7**)

**Table 7:** Influence of Knowledge of dog owners on rabies vaccination uptake in Butaleja TC.

| Knowledge variable | Given score | Vaccinated n(%) | None vaccinated n(%) | cPR (95% CI) | aPR (95% CI) |
| --- | --- | --- | --- | --- | --- |
| **Mode of transmission** |  |  |  |  |  |
| No correct answer | 0 | 11 (68.7) | 5 (31.3) | 1 | 1 |
| One correct answer | 1 | 10 (43.5) | 13 (56.5) | 0.62 (0.31 - 1.22) | 0.55 (0.33 - 1.18) |
| Two correct answers | 2 | 61(62.2) | 37 (37.8) | 1.31 (1.23 - 2.38) | 1.43 (1.32 - 2.44)** |
| Three or more correct answers | 3 | 18 (50.0) | 18 (50.0) | 1.02 (0.66 - 1.40) | 1.11 (0.75 - 1.47) |

| **Importance of vaccination** |  |  |  |  |  |
| --- | --- | --- | --- | --- | --- |
| One correct answer | 1 | 3 (33.3) | 6 (66.7) | 1 | 1 |
| Two correct answers | 2 | 39 (58.2) | 28 (41.8) | 1.92 (1.03 - 3.59) | 1.85 (1.23 - 2.41)** |
| Three or more correct answers | 3 | 58 (59.8) | 39 (40.2) | 2.11 (1.14 - 3.91) | 2.05 (1.10 - 3.80)** |
| **Rabies signs** |  |  |  |  |  |
| No correct answer | 0 | 22 (43.1) | 29 (56.9) | 1 | 1 |
| One correct answer | 1 | 2 (50.0) | 2 (50.0) | 1.35 (0.51 - 3.54) | 1.30 (0.48 - 3.45) |
| Two correct answers | 2 | 16 (61.5) | 10 (38.5) | 1.36 (1.05 - 2.14) | 1.42 (1.21 - 2.51)** |
| Three or more correct answers | 3 | 60 (65.2) | 32 (34.8) | 1.81 (1.27 - 2.55) | 1.73 (1.22 - 2.52)** |

## 4.0 Discussion

This study assessed rabies vaccination uptake among dog owners in Butaleja Town Council, Uganda, and identified factors associated with vaccination decisions at both household and dog population levels. The findings demonstrate that while 57.2% of owners reported vaccinating at least one dog and 81.8% achieved complete household vaccination, overall canine vaccination coverage stood at 48.4%, falling short of the 70% herd immunity threshold recommended for rabies elimination (2, 37). This discrepancy between owner-level uptake and dog-level prevalence underscores the importance of reporting both metrics to avoid misleading interpretations of program performance (38, 39). The observed coverage gap reflects persistent structural and behavioral barriers that require targeted, multi-sectoral interventions to achieve the global “Zero by 30” target (2, 40).

The partial household vaccination pattern identified in this study represents a critical finding with important implications for rabies control programming. While aggregate coverage estimates in Uganda have traditionally been cited at approximately 10% (40, 41), our findings suggest that once owners engage with vaccination services, they are likely to complete vaccination across their entire dog population. This behavioral pattern has been observed in other endemic settings where initial engagement with veterinary services predicts comprehensive household coverage (42, 43). However, the 48.4% coverage level remains insufficient to interrupt transmission, consistent with evidence from sub-Saharan Africa where coverage estimates frequently fall below the 70% threshold despite moderate owner engagement (44, 45).

Socio-demographic factors demonstrated consistent associations with vaccination uptake, aligning with established patterns in rabies-endemic settings. Older dog owners showed higher vaccination likelihoods compared to younger owners aged ≤20 years. This age gradient corresponds with findings from Tanzania and Nigeria where older owners demonstrated greater awareness and perceived responsibility (24, 25).The association between education and vaccination reinforces the role of health literacy in preventive health behaviors (26, 27). Gender differences, with male owners showing lower uptake compared to females, reflect access to resources and decision-making patterns observed elsewhere (28, 46). The positive association with higher income and extended family structures suggests that financial resources and social support networks facilitate vaccination, consistent with findings from Vietnam and the Philippines (47, 48).

Veterinary system factors emerged as critical enablers of vaccination uptake. Awareness of vaccination policies and receipt of vaccination reminders showed strong associations with uptake, highlighting the importance of communication strategies in promoting vaccination adherence (49). Access to veterinary clinics was strongly associated with vaccination, corroborating evidence from Haiti and Tanzania where veterinary infrastructure availability predicts vaccination coverage (45, 47). These findings underscore the need for strengthened veterinary service delivery, particularly in underserved peri-urban areas where access remains limited.

Health system factors similarly were associated with vaccination decisions through multiple pathways. Awareness of vaccination locations and knowledge of post-exposure prophylaxis were associated with increased uptake, suggesting that integrated human and animal health messaging may enhance vaccination programs (37). Community education participation showed particularly strong associations, consistent with evidence from Ghana and Chad demonstrating the effectiveness of community engagement in promoting rabies prevention (50, 51).The association between comfort discussing dog health and vaccination uptake points to the importance of trust and communication in veterinary-client relationships (51).

Dog-related factors revealed important patterns in vaccination behavior. Dogs confined outdoors or roaming freely showed higher vaccination rates than predominantly indoor dogs, likely reflecting perceived rabies exposure risk (24, 26). Dogs used for security purposes were more likely vaccinated, consistent with findings where perceived utility influences preventive care decisions (27, 28). Households with multiple dogs demonstrated higher vaccination rates, potentially due to heightened responsibility and perceived risk among owners with larger dog populations. The lower vaccination likelihood for dogs brought from outside town suggests gaps in vaccination outreach for newly introduced animals, a finding with implications for targeted interventions at points of dog movement and acquisition.

Knowledge emerged as a powerful factor associated with vaccination behavior across multiple dimensions. Owners correctly answering questions about rabies transmission showed progressively higher vaccination likelihood. Similarly, understanding vaccination importance was strongly associated with uptake, with those answering three or more questions correctly showing the highest likelihood. Awareness of rabies signs also predicted vaccination, with owners correctly identifying signs showing greater uptake. These dose-response relationships align with evidence from Tanzania and Bhutan demonstrating that comprehensive knowledge of rabies prevention is associated with vaccination behavior (2, 25, 52).

The public health implications of these findings are substantial for rabies elimination efforts in Uganda and similar endemic settings. First, the coverage gap between owner-level uptake and dog-level prevalence suggests that household-level interventions may be more effective than dog-level approaches, particularly given the high completeness among vaccinating households. Second, the multiple pathways through which socio-demographic, veterinary, health system, dog-related, and knowledge factors influence uptake indicate that single interventions are unlikely to achieve the 70% coverage target. Third, the strong associations with communication, reminders, and community education suggest that demand-side interventions addressing awareness and access may be as important as supply-side improvements in service delivery.

Several strengths of this study merit consideration. Vaccination status verification using available records reduced misclassification bias (53). Stratified sampling ensured representation across the study population (54, 55), and modified Poisson regression controlled for potential confounders (56, 57). However, limitations must be acknowledged. The cross-sectional design precludes causal inference, and self-reported vaccination data may be subject to recall and social desirability bias (58–60). The restriction to Butaleja TC limits generalizability to areas with different socioeconomic and cultural contexts (61). To mitigate these limitations, the questionnaire was pretested in a comparable setting, research assistants received standardized training, and vaccination data were cross-checked with available records where possible (62–64). Clear question design further reduced potential misunderstandings during interviews (65).

Achieving the global “Zero by 30” target for eliminating dog-mediated human rabies hinges on reaching and sustaining the critical 70% vaccination coverage threshold in canine populations (2, 37, 40). However, in many low-resource, endemic settings including Uganda, coverage estimates remain persistently low, often cited at approximately 10% (40, 41). A growing body of evidence from sub-Saharan Africa has identified a recurring set of barriers: limited community awareness, high costs, logistical constraints, and weak veterinary infrastructure (44). This study contributes to this evidence base by shifting the analytical lens from aggregate dog-level vaccination coverage to household-level decision-making, offering a more granular understanding of the behavioral and structural factors associated with vaccination uptake in a peri-urban Ugandan setting. Intensified, multi-pronged approaches are required to address these identified barriers. Outreach should target younger, less educated, and lower-income dog owners using familiar communication channels including local media, community meetings, and peer networks. Expanding access through mobile vaccination teams and convenient service points, alongside reliable reminder systems using radios, Village Health Teams, and community leaders, can close awareness and logistical gaps. Financial incentives, potentially integrated into existing poverty reduction programs such as the Parish Development Model, can address affordability particularly for low-income and younger single owners. Trusted community figures should be engaged to counter myths and promote responsible dog ownership, normalizing vaccination practices. Line ministries including MAAIF and MoH must strengthen risk communication in underserved areas while aligning interventions with dog-keeping purposes to enhance impact. Finally, empowering district veterinary services to regularly monitor and evaluate rabies control efforts is essential for adaptive, data-informed programming.

In conclusion, while this study reveals meaningful progress in rabies vaccination uptake among dog owners in Butaleja Town Council, the dog-level coverage falls below the 70% herd immunity threshold necessary for rabies elimination. Achieving global targets requires sustained, coordinated interventions that address the multiple socio-demographic, knowledge, service delivery, and system factors associated with vaccination uptake. Strengthening veterinary services, increasing community awareness, ensuring vaccine accessibility, and promoting integrated One Health approaches remain essential to advancing rabies elimination in Uganda and similar endemic settings.

## Data Availability

Due to ethical restrictions related to participant confidentiality, the anonymized individual-level dataset cannot be made publicly available. De-identified data supporting the findings of this study are available upon reasonable request from the corresponding author and with approval from the Makerere University School of Public Health Research and Ethics Committee (MakSPH-REC), Institutional Review Board, for researchers who meet the criteria for access to confidential data.

## Author Contributions

SH: Conceptualization, Formal analysis, Investigation, Methodology, Project administration, Resources, Data curation, Validation, Visualization, Writing – original draft, Writing – review & editing.

SK: Conceptualization, Formal analysis, Methodology, Writing – review & editing.

SML: Conceptualization, Writing – review & editing.

DL: Conceptualization, Formal analysis, Methodology, Writing – review & editing.

## 7.0 Acknowledgement

The author(s) humbly express their sincere gratitude to Makerere University School of Public Health for the academic guidance and support extended throughout this work. Appreciation is also extended to staffs of Butaleja district local government for their cooperation, and to the Department of Integrated Epidemiology, Surveillance and Public Health Emergencies, Ministry of Health, Uganda for their invaluable technical input and career guidance.

## Notes

### Competing Interest Statement

The authors have declared no competing interest.

### Author Declarations

This study was approved by the Makerere University School of Public Health Research and Ethics Committee (MakSPH-REC), Institutional Review Board, under Protocol No. 397. Written and verbal informed consent were obtained from all participants prior to enrolment. Participation was voluntary, and all data were anonymized to protect participant confidentiality.

